# N-protein vaccine Convacell® is effective against COVID-19: phase 3, randomized, double-blind, placebo-controlled clinical trial

**DOI:** 10.1101/2024.04.01.24304717

**Authors:** Sevastyan O. Rabdano, Ellina A. Ruzanova, Anastasiya E. Vertyachikh, Valeriya A. Teplykh, Alla B. Emelyanova, German O. Rudakov, Sergei A. Arakelov, Iuliia V. Pletyukhina, Nikita S. Saveliev, Anna A. Lukovenko, Liliya N. Fakhretdinova, Ariana S. Safi, Ekaterina N. Zhirenkina, Irina N. Polyakova, Natalia S. Belozerova, Vladislav V. Klykov, Arina P. Savelieva, Aleksey A. Ekimov, Vadim A. Merkulov, Sergei M. Yudin, Daria S. Kruchko, Igor A. Berzin, Veronika I. Skvortsova, Clinical Trial Group

## Abstract

We have developed a Convacell®, a COVID-19 vaccine based on the conservative viral nucleocapsid (N) protein. The N protein is evolutionary conservative and is abundantly expressed on the surface of infected cells, allowing anti-N immune response generated by Convacell® to rapidly clear infected cells and provide long-lasting protection against COVID-19. Convacell® has been demonstrated to be safe and highly immunogenic, creating immune responses lasting over a year, in phase I/II and IIb clinical trials. Phase IIb clinical trial has also demonstrated that a single dose vaccination regimen with Convacell® is sufficient to provide an immune response.

Here we report the finding of the phase III clinical trial of Convacell®. Two groups of volunteers from Russia have been either vaccinated with a single dose of Convacell® or injected with placebo, and then monitored for incidence of COVID-19 and adverse effects. Anti-N antibody titers at admission were also analyzed, to take into account for potential effects of previous virus encounters.

Disease incidence over 6 months results indicate an overall vaccine efficacy of 85.2% (95% confidence interval: 67.4-93.3%). Additionally, Convacell® has shown a good safety profile. Overall, Convacell® demonstrated highly desirable qualities and good performance as a vaccine and can be considered as valuable COVID-19 preventative measure.

## Introduction

Anti-SARS-CoV-2 vaccines are hailed as the best and most cost-effective treatment for COVID-19 (1), crucial for protecting the vulnerable groups from the disease and lowering its economic and social impact (2–4).

We have developed a COVID-19 vaccine Convacell® based on the full-length nucleocapsid (N) protein of SARS-CoV-2, produced in an *Escherichia coli* recombinant protein platform (5). COVID-19 vaccines based on the N protein have already been described as promising in multiple papers (6–12). Previously, Convacell® has been shown to be effective in protecting from severe disease in preclinical trials (5) and highly immunogenic and safe in phase I, II and IIb clinical trials (13).

Convacell® can be used for vaccination of the general populace. Additionally, Convacell® contains a different target antigen – protein N – and protects via a different dominant immune mechanism, compared to most currently available COVID-19 vaccines, which are based on the spike (S) protein of SARS-CoV-2. As such, we theorize it will be especially useful for people who are unable to generate an anti-S immune response (14–19). Unlike most common vaccines, Convacell® requires only a single dose to achieve protective effectiveness (13) and does not require booster doses to generate an immune response lasting up to a year (13).

In this study, we describe the results of the phase III of Convacell®’s clinical trials, which include assessments of Convacell®’s vaccine efficacy and safety.

## Methods

### Study design

The phase III study (NCT05726084) was planned to be prospective, multicenter, randomized, double-blind and placebo controlled. Study involve comparison between vaccine and placebo groups of volunteers to assess adverse effects (AE) and COVID-19 infection incidence and adherence to protocol. On-site monitoring by research centers ensured that Good Clinical Practices were followed throughout the course of the study. The protocol followed the Helsinki Declaration Guidelines and was firstly evaluated and approved by ethical committee at Ministry of Health of Russian Federation and then by independent onsite ethical committees. The study design included the possibility of preterm conclusion of the study after recruitment of either 33% or 66% of the target number of participants, if the gathered data allowed for conclusive assessment of vaccine efficacy in the vaccinated group greatly exceeding that in the placebo group.

The study recruited volunteers meeting all of the following criteria. Inclusion criteria:

1. Age >18.
2. Willing to sign an informed consent statement to participate in a clinical trial.
3. 18,5 ≤ BMI ≤ 30 kg/m^2^, with body mass between 55 and 100 kg for men and between 45 and 100 kg for women.
4. Verified healthy status: no deviation from reference intervals in the results of standard clinical and laboratory tests.
5. Negative for human immunodeficiency virus (HIV), rapid plasma reagin (RPR), hepatitis B surface antigen (HBsAg), hepatitis C virus RNA (HCVRNA).
6. Haemodynamic and vital parameters within following reference intervals: heart rate 60–90 bpm, respiratory rate under 22 breaths per minute, systolic arterial pressure 100–139 mmHg, diastolic arterial pressure 60–89 mmHg.
7. Willing to keep a self-observation diary and attend control visits.
8. Willing to abstain from alcohol for 14 days before the beginning of the study and until its completion.
9. Willing to abstain from smoking for 48 hours before the beginning of the study and on the admission day.
10. For fertile women: negative pregnancy test and willing to use adequate contraception methods until the completion of the study and for at least two months after vaccination.
11. For fertile men: willing to use adequate contraception methods until the completion of the study or past vasectomy with confirmed azoospermia, partner willing to use at least 90% effective contraception methods or past tubal ligation or menopausal for at least 2 years.

### Procedures

The volunteers in the study received one dose of a recombinant subunit COVID-19 vaccine based on the nucleocapsid protein of SARS-CoV-2, Convacell®, in the form of emulsion for intramuscular injection. The vaccine used in this study was produced according to the GMP by the Saint Petersburg Scientific Research Institute of Vaccines and Serums (SPbSRIVS). The one-dose vaccination regimen was chosen following the results of the earlier IIb study (13).

Each 0.5 ml dose of Convacell® contains 50 μg of recombinant SARS-CoV-2 nucleocapsid protein as the main active ingredient. Supplementary ingredients are 5 mg of (±)-α-tocopherol, 15 mg of squalane and 5 mg of polysorbate 80 in form of nanoemulsion.

The placebo formulation used in the study was identical to the vaccine formulation, with the exception of containing no SARS-CoV-2 nucleocapsid protein.

Participants’ sera was collected on screening and during unplanned visits. To obtain volunteer sera, volunteers’ blood was collected into 6 ml vacuum tubes containing K2 EDTA as anticoagulant. The blood was processed to sera using centrifugation. The quantities of specific anti-N IgG antibodies in volunteer sera were assessed via the AdviseDx SARS-CoV-2 IgG II chemiluminescent microparticle immunoassay (Abbott Laboratories, Chicago, United States) and Anti-N IgG ELISA kit (St. Petersburg Research Institute of Epidemiology and microbiology, Russia). The standard manufactuter’s protocol was followed.

### Randomization and masking

Both the placebo and the vaccine doses appeared identical on external examination: as opaque milky-white suspension. Placebo and vaccine doses were supplied in visually identical vials containing no markings as to the nature of the suspension, except a numerical id-tag that was deliberately meaningless to participating volunteers and their physicians. Common practices included supplying both placebo and vaccine dose vials intermixed within same shipping containers. Staff that were unaware of assignments of subjects carried out data management and statistical analyses.

### Outcomes

The main endpoint of this phase III study was the frequency of PCR-confirmed COVID-19 infections among the participants, including asymptomatic infections, after 15 days since vaccination and until 6 months after vaccination. The secondary endpoints were: 1) the frequency of COVID-19, here meaning a SARS-CoV-2 infection coupled with at least one common COVID-19 symptom (fever/chills, cough, shortness of breath or difficulty breathing, fatigue, muscle or body pain, headache, loss of taste or smell, sore throat, congestion or runny nose, nausea or vomiting, diarrhea) after 15 days since vaccination and until 6 months after vaccination; 2) the titers of anti-N IgG present at volunteer admission into the study and 3) the frequency of local and systemic AEs observed during the first 28 days since vaccination. AE were determined as disturbances in the tested vital parameters; or disorders that arose during the course of the study and were detected as the result of assessment by a professional physician. All AE that were observed during the course of the safety study were recorded, regardless of their putative association with the administration of the studied vaccine formulation. Severe AE (SAE) were defined as any AE that led to hospitalization of the volunteer and/or required immediate medical intervention, and/or led to the volunteer’s death. Volunteers were instructed to visit the hospital if they considered themselves to be having COVID-19 symptoms, during this visit, volunteers were examined by professional physicians and all potential COVID-19 symptoms as well as the infection itself were recorded as AEs.

### Statistical analysis

Formal sample size calculations were carried out via a validated copy of PASS 2021 software, version 21.0.5 (NCSS Statistical Software, United States), itself based on the works of O’Hagan, Stevens and Campbell in sample size statistics (20).

The mode used was “superiority by a margin test for vaccine efficacy using the ratio of two proportions.” Vaccine efficacy (VE) for the placebo group was set to ≤ 30%, VE for the vaccinated group was set to 70%, infection incidence over a 6 month period was set to 0.6%, confidence interval was set to 95%, dropout rate during the study were set to 10%. Ratio between the placebo group and the vaccinated group was set to be 1:2.

The results indicated that 9505 participants were required to be included into the vaccinated group and 4753 into the vaccinated group, or 14258 participants were required in total. The total number of screened individuals to reach the needed number of participants was 17600.

Due to the unknown future incidence of infection in the target population over the course of the study, during the planning stage, it was decided in the protocol to recruit participants in successive stages. Initially, 33% of the participants were enrolled, treated and observed. Due to higher incidence of COVID-19 in the sample population compared to the one assumed during the sample size calculation, the next stages of enrollment were judged to not be necessary to meet the planned endpoints and the clinical trial was concluded.

## Results

Enrollment of volunteers started on May 18, 2023 and ended on August 9, 2023. Screening, randomization and populations details are given in Fig. 1. The observation for the last enrolled volunteer ended on February 11, 2024. For the “intention to treatment” efficacy population 8 cases of PCR confirmed COVID-19 were detected in the vaccine group, while in placebo group there were 27 cases. Vaccine efficacy (Fig. 2) assessment results indicate a high vaccine efficacy of 85.2% (95% confidence interval: 67.4-93.3%), with the attack rate in the vaccinated group being 5.78 times lower than in the placebo group (0.23% vs 1.55%). The primary endpoint regarding vaccine efficacy has been conclusively reached by the available data.

**Fig 1:**
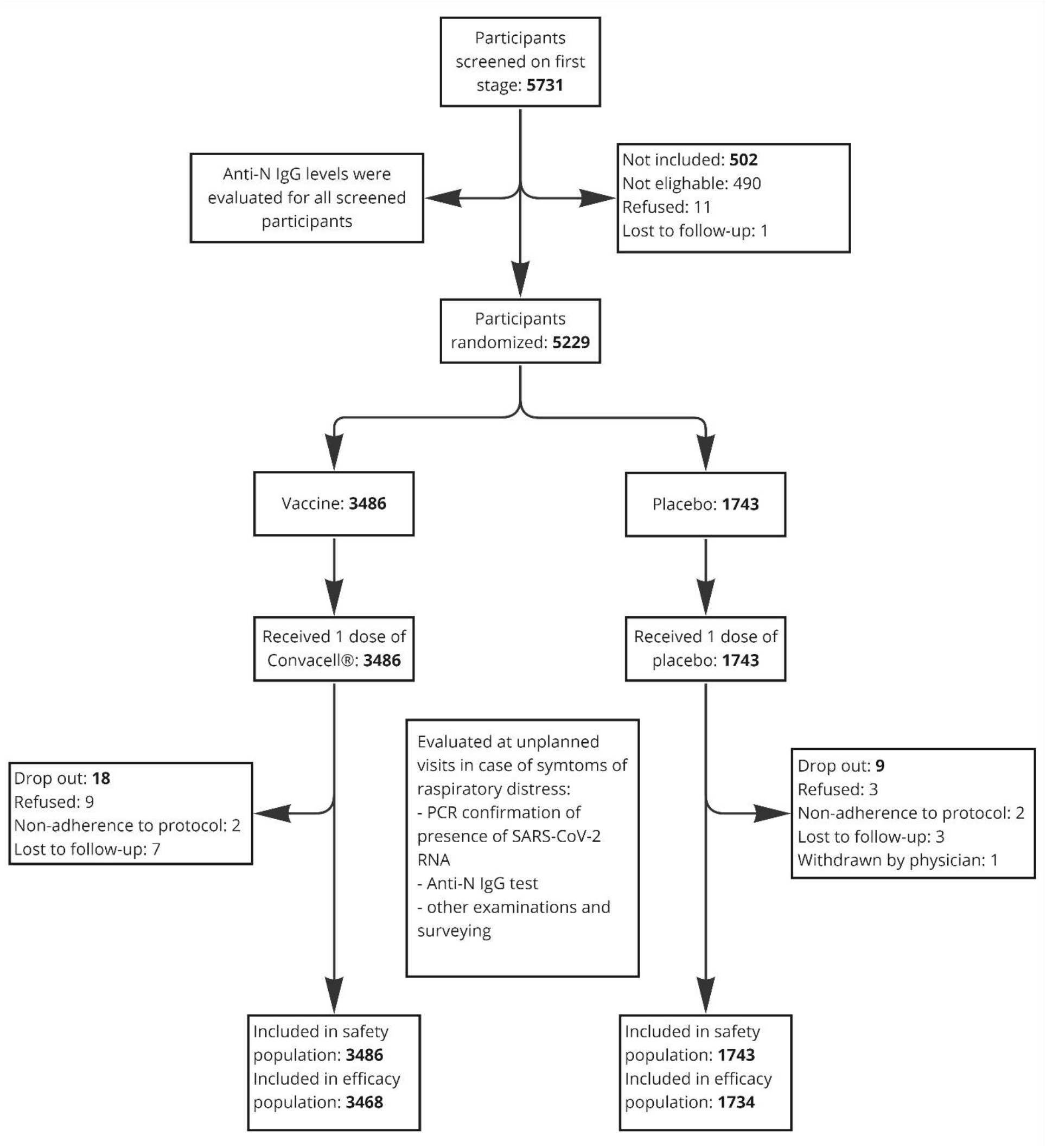
Schematic description of the clinical trial.

**Figure 2:**
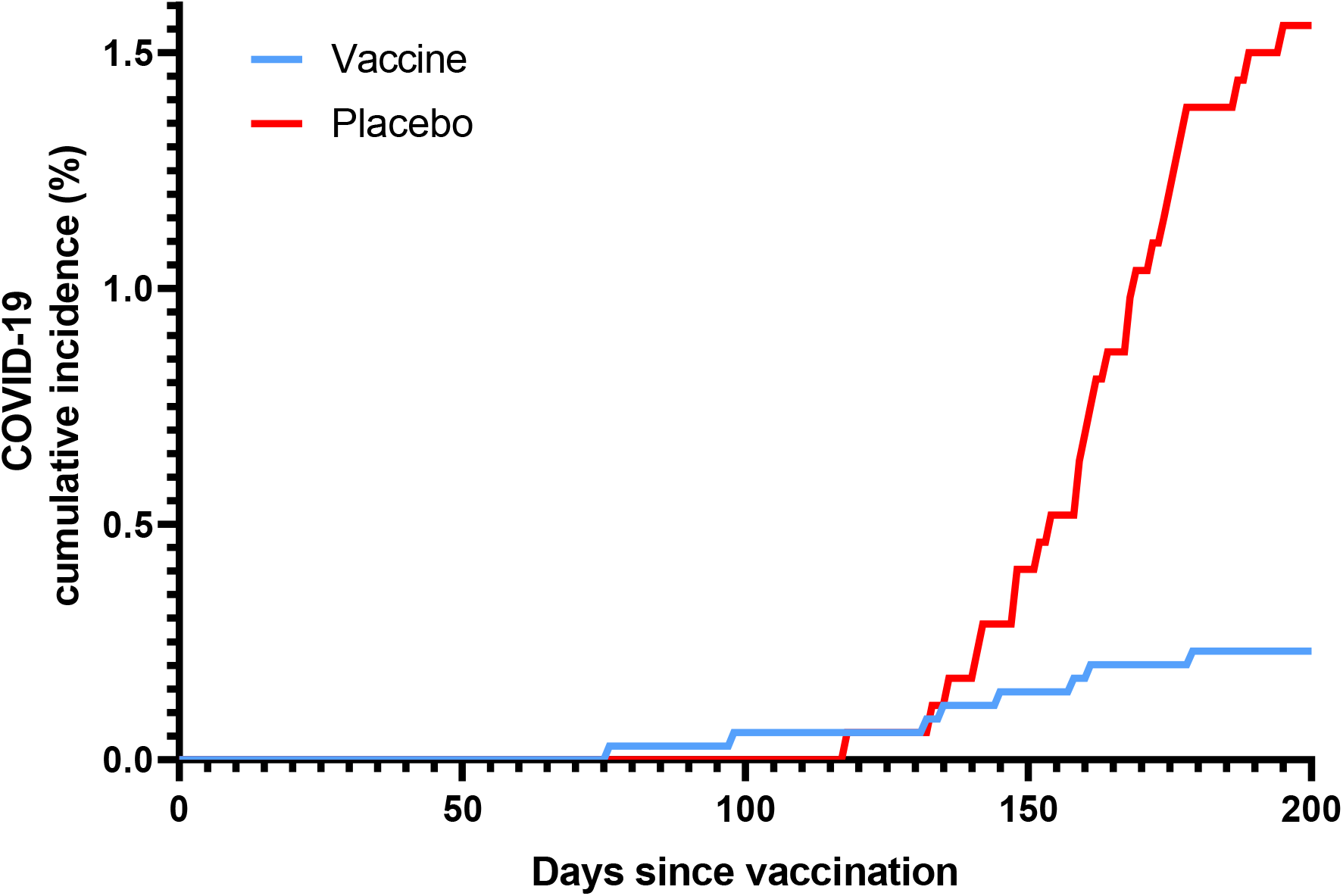
Vaccine efficacy graph showing cumulative incidence rate of COVID-19 among two groups of volunteers normalized for days after vaccination.

Analysis of anti-N antibody titers revealed that the fraction of participants with detectable anti-N antibody titers was 29.26% in the vaccinated group and 30.12% in the placebo group. As such, no major differences in rate of COVID-19 naivety between the two groups were discovered and the secondary endpoint regarding the titers of anti-N antibodies on randomization in participants has been reached.

Safety assessment of the vaccine demonstrated high safety throughout, with most commonly observed adverse effects (Fig. 3) being mild reaction site conditions or mild systemic disturbances. Notably, vaccinated group had higher incidence of adverse effects compared to the placebo group only with regards to reaction site conditions, malaise, gastrointestinal pain and hyperhidrosis.

**Figure 3:**
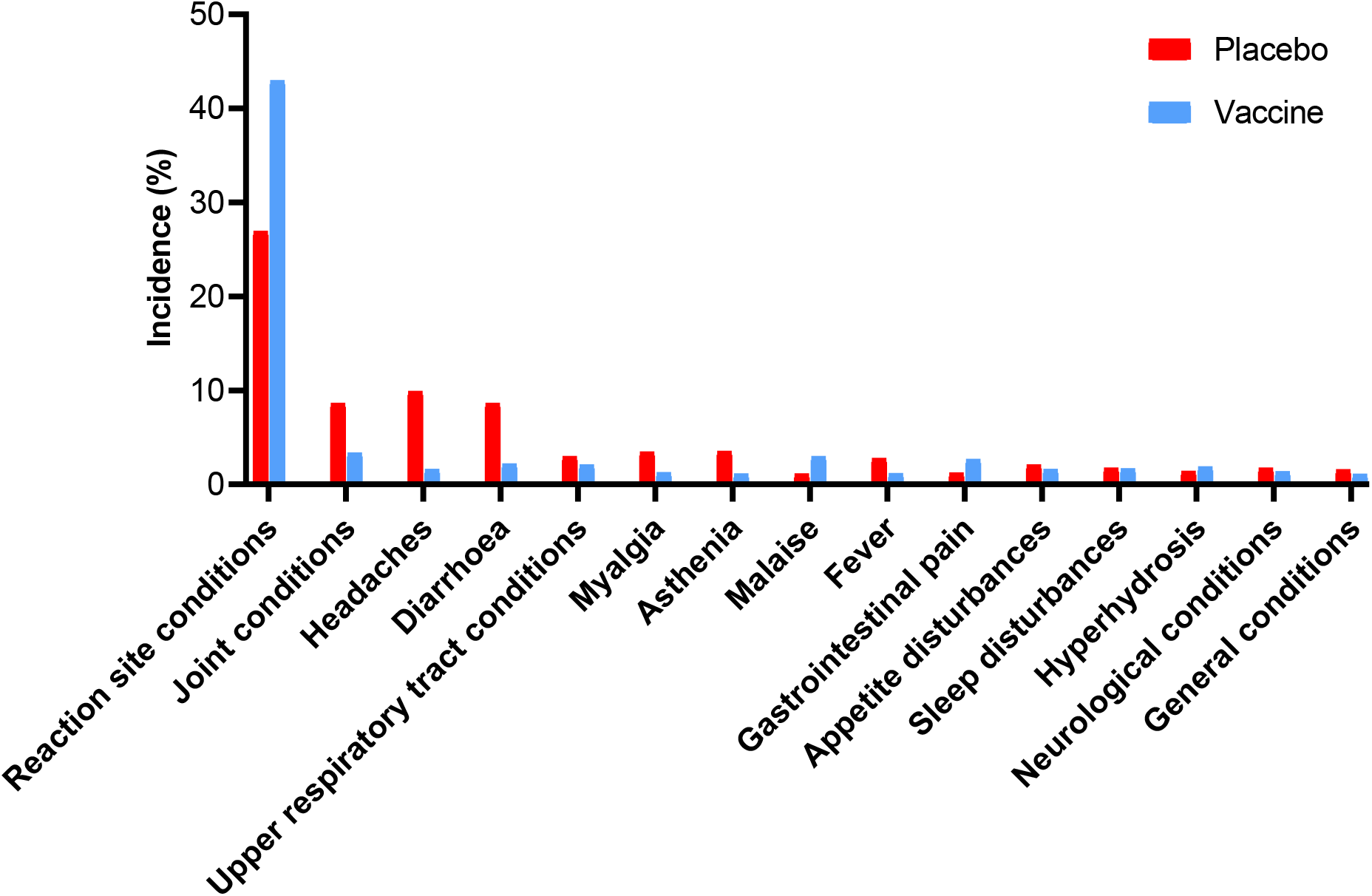
Incidence rates of adverse effects affecting more than 1% of participants, for both groups. Only adverse effects that has possible, probable, and definite causality are counted in figure.

## Discussion

All the planned endpoints have been successfully reached by the trial, which allows for the conclusion that, based on the obtained data, Convacell® has a high vaccine efficacy of 85.2%. Convacell®’s protection is evidently long lasting, given the lack of convergence between the placebo and vaccinated groups’ COVID-19 incidence rates over the 180∼195 days of observation in this study. Based on the immune response longevity data obtained in the previous phases of Convacell®’s study (13), we can reasonably assume that Convacell®’s protection should last at least a year, given that vaccinated individuals are positive for anti-N antibodies for at least a year after vaccination.

The usage of the internal N protein as the main antigen in Convacell® confers on it a number of advantages. The N protein is notably conservative (7,8,21–24), which confers onto Convacell®-generated immune responses both longevity (5) and broad cross-reactivity among SARS-CoV-2 variants (25). The internal nature of the N protein in mature virions does not negatively affect Convacell®’s protectivity, as the N protein is highly expressed in infected cells (26,27) and exposed on their membranes (28,29), which allows such cells to be targeted and eliminated by cytotoxic T-cell (30–32) and natural killer (NK) cell action (33–35). Currently, highly efficient infected cell clearance is theorized to be the main protective mechanism of Convacell®-generated immunity, based on the antibody-dependent NK cell activation data obtained in phase II of Convacell®’s clinical trials (13).

Convacell®’s recombinant *E. coli* protein platform confers onto it a very desirable safety profile. The most frequent adverse effects among vaccinated and placebo groups were local injection site reactions, with the rate of most frequent adverse effects – localized injection site reactions – observed in the vaccination group being considerably lower than that reported in the clinical trials of common COVID-19 vector vaccines. Overall, Convacell® demonstrated highly desirable qualities and good performance as a vaccine and can be considered as valuable COVID-19 preventative measure.

## Supporting information

Clinical Trial Group

## Data Availability

All data produced in the present study are available upon reasonable request to the authors.

## References

1. Miao G, Chen Z, Cao H, Wu W, Chu X, Liu H, et al. From Immunogen to COVID-19 vaccines: Prospects for the post-pandemic era. Biomed Pharmacother. 2023 Feb;158:114208.

2. Yong SJ. Long COVID or post-COVID-19 syndrome: putative pathophysiology, risk factors, and treatments. Infect Dis (Lond). 2021 Oct;53(10):737–54.

3. Raveendran AV, Jayadevan R, Sashidharan S. Long COVID: An overview. Diabetes Metab Syndr. 2021;15(3):869–75.

4. Bloom N, Bunn P, Mizen P, Smietanka P, Thwaites G. The Impact of COVID-19 on Productivity. The Review of Economics and Statistics. 2023 Feb 6;1–45.

5. Rabdano SO, Ruzanova EA, Pletyukhina IV, Saveliev NS, Kryshen KL, Katelnikova AE, et al. Immunogenicity and In Vivo Protective Effects of Recombinant Nucleocapsid-Based SARS-CoV-2 Vaccine Convacell®. Vaccines (Basel). 2023 Apr 20;11(4):874.

6. Bai Z, Cao Y, Liu W, Li J. The SARS-CoV-2 Nucleocapsid Protein and Its Role in Viral Structure, Biological Functions, and a Potential Target for Drug or Vaccine Mitigation. Viruses. 2021 Jun 10;13(6):1115.

7. Thura M, Sng JXE, Ang KH, Li J, Gupta A, Hong JM, et al. Targeting intra-viral conserved nucleocapsid (N) proteins as novel vaccines against SARS-CoVs. Bioscience Reports. 2021 Sep 30;41(9):BSR20211491.

8. Dutta NK, Mazumdar K, Gordy JT. The Nucleocapsid Protein of SARS–CoV-2: a Target for Vaccine Development. Dutch RE, editor. J Virol. 2020 Jun 16;94(13):e00647–20, /jvi/94/13/JVI.00647-20.atom.

9. Tilocca B, Soggiu A, Sanguinetti M, Musella V, Britti D, Bonizzi L, et al. Comparative computational analysis of SARS-CoV-2 nucleocapsid protein epitopes in taxonomically related coronaviruses. Microbes and Infection. 2020 May;22(4–5):188–94.

10. Sieling P, King T, Wong R, Nguyen A, Wnuk K, Gabitzsch E, et al. Prime hAd5 Spike + Nucleocapsid Vaccination Induces Ten-Fold Increases in Mean T-Cell Responses in Phase 1 Subjects that are Sustained Against Spike Variants [Internet]. Allergy and Immunology; 2021 Apr [cited 2022 Nov 14]. Available from: http://medrxiv.org/lookup/doi/10.1101/2021.04.05.21254940

11. Matchett WE, Joag V, Stolley JM, Shepherd FK, Quarnstrom CF, Mickelson CK, et al. Cutting Edge: Nucleocapsid Vaccine Elicits Spike-Independent SARS-CoV-2 Protective Immunity. JI. 2021 Jul 15;207(2):376–9.

12. Van Elslande J, Oyaert M, Ailliet S, Van Ranst M, Lorent N, Vande Weygaerde Y, et al. Longitudinal follow-up of IgG anti-nucleocapsid antibodies in SARS-CoV-2 infected patients up to eight months after infection. Journal of Clinical Virology. 2021 Mar;136:104765.

13. Rabdano S, Ruzanova E, Makarov D, Vertyachikh A, Teplykh V, Rudakov G, et al. Safety and Immunogenicity of the Convacell® Recombinant N Protein COVID-19 Vaccine. Vaccines. 2024 Jan 19;12(1):100.

14. Gattinger P, Kratzer B, Tulaeva I, Niespodziana K, Ohradanova-Repic A, Gebetsberger L, et al. Vaccine based on folded receptor binding domain-PreS fusion protein with potential to induce sterilizing immunity to SARS-CoV-2 variants. Allergy. 2022 Aug;77(8):2431–45.

15. Wagner A, Garner-Spitzer E, Schötta AM, Orola M, Wessely A, Zwazl I, et al. SARS-CoV-2-mRNA Booster Vaccination Reverses Non-Responsiveness and Early Antibody Waning in Immunocompromised Patients – A Phase Four Study Comparing Immune Responses in Patients With Solid Cancers, Multiple Myeloma and Inflammatory Bowel Disease. Front Immunol. 2022 May 12;13:889138.

16. Mair MJ, Mitterer M, Gattinger P, Berger JM, Trutschnig W, Bathke AC, et al. Enhanced SARS-CoV-2 breakthrough infections in patients with hematologic and solid cancers due to Omicron. Cancer Cell. 2022 May;40(5):444–6.

17. Francis AI, Ghany S, Gilkes T, Umakanthan S. Review of COVID-19 vaccine subtypes, efficacy and geographical distributions. Postgrad Med J. 2022 May;98(1159):389–94.

18. Garcia P, Anand S, Han J, Montez-Rath M, Sun S, Shang T, et al. COVID19 vaccine type and humoral immune response in patients receiving dialysis. medRxiv. 2021 Aug 4;2021.08.02.21261516.

19. Ferri C, Ursini F, Gragnani L, Raimondo V, Giuggioli D, Foti R, et al. Impaired immunogenicity to COVID-19 vaccines in autoimmune systemic diseases. High prevalence of non-response in different patients’ subgroups. J Autoimmun. 2021 Dec;125:102744.

20. O’Hagan DT, Ott GS, Nest GV, Rappuoli R, Giudice GD. The history of MF59 ® adjuvant: a phoenix that arose from the ashes. Expert Review of Vaccines. 2013 Jan;12(1):13–30.

21. Yang H, Rao Z. Structural biology of SARS-CoV-2 and implications for therapeutic development. Nat Rev Microbiol. 2021 Nov;19(11):685–700.

22. Dai L, Gao GF. Viral targets for vaccines against COVID-19. Nat Rev Immunol. 2021 Feb;21(2):73–82.

23. Pack SM, Peters PJ. SARS-CoV-2-Specific Vaccine Candidates; the Contribution of Structural Vaccinology. Vaccines (Basel). 2022 Feb 3;10(2):236.

24. Yewdell JW. Antigenic drift: Understanding COVID-19. Immunity. 2021 Dec 14;54(12):2681–7.

25. Rabdano S, Mukhin V, Makarov V, Rudakov G, Ruzanova E, Arakelov S, et al. N protein based vaccine against SARS-CoV-2 produces a strong T cell immune response to N Protein of novel strains. MES. 2022 Sep;(2022(3)).

26. Zhang XY, Guo J, Wan X, Zhou JG, Jin WP, Lu J, et al. Biochemical and antigenic characterization of the structural proteins and their post-translational modifications in purified SARS-CoV-2 virions of an inactivated vaccine candidate. Emerging Microbes & Infections. 2020 Jan 1;9(1):2653–62.

27. Yu S, Wei Y, Liang H, Ji W, Chang Z, Xie S, et al. Comparison of Physical and Biochemical Characterizations of SARS-CoV-2 Inactivated by Different Treatments. Viruses. 2022 Aug 31;14(9):1938.

28. López-Muñoz AD, Kosik I, Holly J, Yewdell JW. Cell surface SARS-CoV-2 nucleocapsid protein modulates innate and adaptive immunity. Science Advances. 8(31):eabp9770.

29. Fielding CA, Sabberwal P, Williamson JC, Greenwood EJ, Crozier TW, Zelek W, et al. SARS-CoV-2 host-shutoff impacts innate NK cell functions, but antibody-dependent NK activity is strongly activated through non-spike antibodies. eLife. 2022 May 19;11:e74489.

30. Peng Y, Felce SL, Dong D, Penkava F, Mentzer AJ, Yao X, et al. An immunodominant NP105-113-B*07:02 cytotoxic T cell response controls viral replication and is associated with less severe COVID-19 disease. Nat Immunol. 2022 Jan;23(1):50–61.

31. Grifoni A, Weiskopf D, Ramirez SI, Mateus J, Dan JM, Moderbacher CR, et al. Targets of T Cell Responses to SARS-CoV-2 Coronavirus in Humans with COVID-19 Disease and Unexposed Individuals. Cell. 2020 Jun 25;181(7):1489–1501.e15.

32. Peng H, Yang L tao, Wang L yun, Li J, Huang J, Lu Z qiang, et al. Long-lived memory T lymphocyte responses against SARS coronavirus nucleocapsid protein in SARS-recovered patients. Virology. 2006 Aug 1;351(2):466–75.

33. Tso FY, Lidenge SJ, Poppe LK, Peña PB, Privatt SR, Bennett SJ, et al. Presence of antibody-dependent cellular cytotoxicity (ADCC) against SARS-CoV-2 in COVID-19 plasma. PLoS One. 2021;16(3):e0247640.

34. Hagemann K, Riecken K, Jung JM, Hildebrandt H, Menzel S, Bunders MJ, et al. Natural killer cell-mediated ADCC in SARS-CoV-2-infected individuals and vaccine recipients. Eur J Immunol. 2022 Aug;52(8):1297–307.

35. Yu Y, Wang M, Zhang X, Li S, Lu Q, Zeng H, et al. Antibody-dependent cellular cytotoxicity response to SARS-CoV-2 in COVID-19 patients. Signal Transduct Target Ther. 2021 Sep 24;6(1):346.

