## Supplementary material for "N-protein vaccine Convacell® is effective against COVID-19: phase 3, randomized, double-blind, placebo-controlled clinical trial": Clinical Trial Group

| Name | Affiliation | Email |
| --- | --- | --- |
| Zakharov Konstantin Anatolyevich | Limited Liability Company "Scientific Research Center Eco-safety", legal address: 196143, St. Petersburg, Yuri Gagarin Ave., 65 | |
| Vasiliuk Vasily Bogdanovich | Limited Liability Company "Scientific Research Center Eco-safety", legal address: 196143, St. Petersburg, Yuri Gagarin Ave., 65 | |
| Vasilevskaya Olga Albertovna | Limited Liability Company "Medical Diagnostic Center", legal address: 150000, Yaroslavl, Ushinsky str., 4b | |
| Alpenidze Diana Nodarievna | St. Petersburg State Budgetary Healthcare Institution "City polyclinic No. 117", legal address: 194358, St. Petersburg, Simonov str., 5, building 1, lit. A | |
| Mullagalieva Ekaterina Andreevna | State Autonomous Healthcare Institution "Engels City Clinical Hospital No. 1", legal address: 6 Vesennaya str., Engels, 413116 | |
| Gamova Inna Valeryevna | Limited Liability Company "DNA Research Center", legal address: 280 Rakhov str., Saratov, 410005 | |
| Bouchard Maxim Osmanovich | Limited Liability Company "Medical services", legal address: 194356, St. Petersburg, Vyborg highway, house 40 litera A. | |
| Tomaev Uruzmag Mairbekovich | Limited Liability Company "Firm ORIS"<br>legal address: 117321, Moscow, Trade Union str., 154, building 1 | |
| Bunkova Elena Borisovna | Private institution educational organization of higher education "Medical University "Reaviz"<br>legal address: 443001, Samara, Chapaevskaya str. 227 | |
| Evsina Maria Gennadievna | State Autonomous Healthcare Institution of the Sverdlovsk region "Aramil City Hospital"<br>legal address: 624002, Sverdlovsk region, Aramil, Sadovaya str., 10 | |
| Bakhtina Natalia Vladimirovna | State regional budgetary healthcare Institution "Murmansk Regional Clinical Hospital named after P.A. Bayandin"<br>legal address: 183032, Murmansk, Akademika Pavlova str., 6, building 3 | |
| Shapovalova Yulia Sergeevna | Private healthcare institution "Clinical Hospital "Russian Railways-Medicine" Chelyabinsk""<br>legal address: 41, Zwilling str., Chelyabinsk, 454091 | |
| Saenko Darya Viktorovna | Limited Liability Company "Ultrasound 4D Clinic"<br>legal address: 357502, Pyatigorsk, Kuznechnaya str., No. 26 | |
| Leontieva Marina Alexandrovna | Limited Liability Company "Clinic Zvezdnaya"<br>legal address: 196158, St. Petersburg, Moskovskoe sh., house 5, letter A, room 9-N | |
| Shkarbul Dmitry Yurievich | Limited liability Company "Clinic Zvezdnaya"<br>legal address: 196158, St. Petersburg, Moskovskoe shosse, 5, litera A, room 9-N. | |
| Teplykh Svetlana Valeryevna | Limited Liability Company "Professorial Clinic"<br>legal address: 614070, Perm, Druzhby str., 15 A | |
| Chizhov Danila Alexandrovich | St. Petersburg State Budgetary Healthcare Institution "City Polyclinic No. 106", legal address: 198328, St. Petersburg, Richard Sorge str., 1, lit. A, actual address: 198206, St. Petersburg, Admiral Cherokov, 12, p. 1 | |
| Panevina Vera Sergeevna | St. Petersburg State Budgetary Healthcare Institution "City Polyclinic No. 4", 199178, St. Petersburg, V.O., Bolshoy ave., 59, lit. A | |
| Romanenkov Andrey Andreevich | Limited Liability Company "North-Western Medical Center" (LLC "SZMC"), legal address: 191144, St. Petersburg, vn.ter.g. Smolninskoye municipal district, Moiseenko str., 5, letter A, room. 2nd office 12. The address of the CI: 194355, St. Petersburg, Prospekt Prosveshcheniya, 14 K.4, letter A, room 30-N. | |
| Frolov Alexey Vyacheslavovich- | 445846, Samara region, Tolyatti, 40 let Pobedy str., 51B, 445846, Samara region, Tolyatti, 40 let Pobedy str., 51A | |
| Astankovich Alesya Vladimirovna | Limited Liability Company Sphere-Med, 197342, St. Petersburg, embankment of the Black River, house 41, building 2, letter B | |
